# Rare Dysfunctional Complement Factor I Genetic Variants and Progression to Advanced Age-Related Macular Degeneration

**DOI:** 10.1101/2022.12.20.22283695

**Authors:** Johanna M. Seddon, Bernard Rosner, Dikha De, Tianxiao Huan, Anuja Java, John Atkinson

**Author notes:** Corresponding Author*, Address for Reprints: 55 Lake Avenue North, S3-119, Department of Ophthalmology and Visual Sciences, University of Massachusetts Chan Medical School, Worcester, MA, 01655, USA. **Meeting Presentations:** Association for Research in Vision and Ophthalmology Meeting, May 2022; American Ophthalmological Society Annual Meeting, May 2022; American Society of Retina Specialists, July 2022.

## Abstract

**PURPOSE:** To evaluate associations between rare dysfunctional Complement Factor I (*CFI*) genetic variant status and progression to advanced age-related macular degeneration (AAMD), geographic atrophy (GA) and neovascular disease (NV).

**DESIGN:** Prospective, longitudinal study

**PARTICIPANTS:** Patients aged 55-80 years at baseline identifying as white with non-advanced AMD in one or both eyes at baseline were included. Follow-up grades were assigned as early, intermediate, or AAMD (GA or NV). *CFI* variants were categorized using genotyping and sequencing platforms.

**METHODS:** Analyses were performed using the Seddon Longitudinal Cohort Study (N = 2116 subjects, 3901 eyes, mean follow-up 8.3 years) and the Age-Related Eye Disease Study (N = 2837 subjects, 5200 eyes, mean follow-up 9.2 years). *CFI* rare variants associated with low serum factor I (FI) protein levels and decreased FI function (type 1), other AMD genetic variants, demographic, behavioral, and ocular factors were evaluated. Generalized Estimating Equations methods were used to assess the association between *CFI* rare variants and progression, independent of other genetic variants and covariates.

**MAIN OUTCOME:** Progression to AAMD, GA, or NV.

**RESULTS:** In the prospective cohort of 4953 subjects (9101 eyes with non-advanced AMD at baseline), 1% were type 1 rare *CFI* carriers. Over 12 years, progression to AAMD was 44% for carriers and 20% for non-carriers (P <0.001) while 30% of carriers versus 10% of non-carriers progressed to GA (P < 0.001); 18% of carriers compared to 11% of non-carriers progressed to NV (P = 0.049). *CFI* carriers were more likely to have a family history of AMD (P for trend = 0.035) and a higher baseline AMD grade (P<0.001). After adjusting for all covariates, *CFI* carrier status was associated with progression to GA (OR = 1.91; 95% CI = 1.03, 3.52) but not NV (OR = 0.96). Higher BMI was associated with progression among *CFI* carriers (BMI ≥25 vs <25; OR = 5.8; 95% CI 1.5, 22.3), but not for non-carriers (OR = 1.1; 95% CI = 0.9, 1.3), with P_interaction = 0.011.

**CONCLUSIONS:** Results suggest that carriers of rare dysfunctional type 1 *CFI* variants are at higher risk for progression to AAMD with GA.

**PRÉCIS:** **Rare dysfunctional Complement Factor I (*CFI*) genetic variants are associated with a family history of AMD and increased risk of progression to advanced dry age-related macular degeneration (AMD) with geographic atrophy**.

## INTRODUCTION

Age-related macular degeneration (AMD) has a complex etiology and remains a significant public health problem despite recent advances in treatments. ^1–4^ Patients with neovascular macular degeneration (NV) disease may have residual visual impairment after treatment with intravitreal injections, due to varying degrees of choroidal and retinal atrophy and scarring. The advanced dry form with geographic macular atrophy (GA) is not treatable currently but many clinical trials with promising therapies are underway.

AMD confers a significant individual and societal burden and can lead to loss of independence, increased utilization of health resources and an adverse impact on quality of life.^2–4^ The prevalence is increasing as the proportion of our elderly population rises, and the number of people with AMD is expected to be 196 million in 2020, increasing to 288 million by 2040.^2^ Prevention of AMD and delay in progression to visual loss are therefore key public health challenges.

A composite set of genetic, demographic, environmental and ocular variables can predict with relatively high probability which individuals are at a greater risk of progression to advanced AMD (AAMD).^1,5,6^ Among individuals with the same baseline macular status, a higher genetic burden was associated with an increased likelihood of progression to advanced disease and visual loss.^5,6^ The genetic predictors include both common variants with low to moderate impact and rare variants with higher impact and more defined biologic mechanisms. The rare variants to date are primarily in the complement pathway, including complement factor H (*CFH)*, complement component 3 (*C3)*, complement factor I (*CFI)* and complement component 9 (*C9)*.^7–16^ In patients carrying complement variants, a given retinal injury or damage may lead to excessive complement activation and damage to the retina, retinal pigment epithelium and choroidal vasculature causing acceleration of the disease process.^11^ Furthermore, individuals with rare genetic variants in these genes are more likely to progress and have advanced disease at an earlier age, and individuals with signs of AMD at a younger age are more likely to carry these variants.^6–13^

*CFI* on chromosome 4q25 encodes a serine protease which is a key inhibitor of the complement system. This protease is responsible for cleaving and inactivating C4b and C3b to downregulate the complement system.^17^ This regulatory activity requires a cofactor protein such as Factor H (FH), C4 Binding Protein (C4BP), membrane cofactor protein (MCP; CD46) or complement receptor 1 (CR1; CD35). Factor I (FI) then cleaves and thereby prevents these two proteins from participating in complement activation pathways.^18,19^

Our team first reported the association between the common variant near *CFI* and AMD in 2009, ^20^ and subsequently identified 59 rare genetic variants in this gene that were related to AMD.^13^ In 2015, we demonstrated that individuals with AAMD and rare *CFI* variants commonly have low serum FI levels, and are a substantial risk factor for AMD.^11^ These results were confirmed in an independent UK cohort in 2020.^21^ We proposed that this group of patients with both a rare *CFI* variant and low serum FI is most likely to benefit from complement inhibitory therapy, especially supplementation with FI.^11^

Since about 30-50% of the rare *CFI* variants are not associated with reduced serum antigenic levels, we also evaluated the function of the rare variants in this gene using a serum assay based on the degradation of C3b to iC3b and demonstrated that they can be categorized into three types **(eTable 1 – online only)**. Type 1 variants lead to low serum FI antigenic levels and a corresponding decrease in FI function.^14,21^ There were twenty-three Type 1 variants identified in our AMD cohort which are shown in **Figure 1**. Type 2 variants demonstrated normal serum FI antigenic levels but reduced functional activity. Variants not belonging to Types 1 and 2 were characterized as Type 3 since they demonstrated normal antigenic levels and a more modest decrease in function (less defective than the Type 2 variants but not equivalent in regulatory activity to wild type FI). ^22^

**Figure 1.**
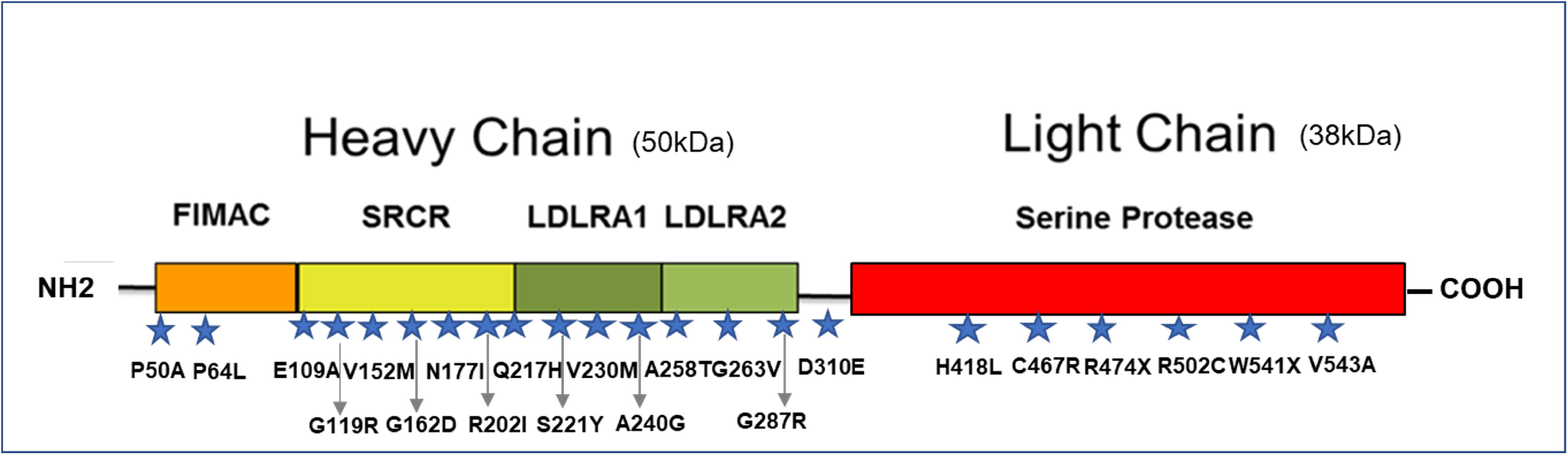
Linear structure of Factor I (FI). Factor I is a multi-domain glycoprotein composed of a heavy chain (50 kDa) and a light chain (38 kDa) held together by a disulfide bond. The heavy chain is composed, from the N-terminal, by the FI Membrane Attack Complex (FIMAC) domain, a Scavenger Receptor Cysteine Rich (SRCR) domain and two Low Density Lipoprotein Receptor class A (LDLRA) domains. The light chain hosts the serine protease domain. There were 23 Type 1 variants identified in our AMD cohort and 22 of them are depicted by blue stars. The variant IVS11+1G>C is an intronic variant and therefore is not shown in the figure.

Although we have shown that Type 1 variants are associated with advanced AMD,^11,14^ to date there is no information about the impact of these rare variants in *CFI* on transition from early or intermediate stages to advanced types of AMD. We, therefore, conducted prospective analyses to determine whether carriers of these variants that allow for amplification of the immune response were at higher risk over time of progressing to advanced AMD. We also assessed whether carriers of types 2 and 3 variants were associated with disease progression compared with non-carriers, despite the lower functional impact of these variants.

## METHODS

### STUDY COHORTS

Details of the AREDS have been previously reported.^23^ The clinical trial was designed to evaluate the effect of antioxidant and mineral supplements on AMD and cataract risk, and risk factors for progression to advanced stages of AMD were evaluated. Participants were aged 55 to 80 years old at baseline. A total of 2837 people (5200 eyes) who were white and had non-advanced AMD in one or both eyes at baseline were included Mean age was 68.8 years with a standard deviation of 4.9 years and mean follow-up time was 9.2 years with an interquartile range of 8.0 to 11.0 years. The Seddon Longitudinal Cohort (SLCS) is a large independent AMD cohort which began in 1985 (J.M.S, Principal Investigator).^1,5^ Participants were enrolled in ongoing epidemiologic and genetic studies of AMD including a registry and biorepository of genetic and other biologic samples, as well as prospective assessment of progression and risk factors for disease. Participants were derived from clinic populations, family and nationwide referrals. The follow-up time was truncated to 12 years, baseline age was restricted to 55-80 to match the follow-up time and age of the AREDS cohort, and individuals who had non-advanced AMD in one or both eyes at baseline were included (N = 2456). Individuals with missing covariates and genes were excluded (N = 340), leaving 2116 people and 3901 eyes in the analysis dataset. Mean age was 68.9 years with a standard deviation of 6.6 years and mean follow-up time was 8.3 years with an interquartile range of 5.0 to 12.0 years. Informed consent was obtained from participants, the research adhered to the tenets of the Declaration of Helsinki and was performed under approved institutional review board protocols.

### DEMOGRAPHIC, LIFESTYLE, OCULAR AND GENETIC FACTORS

Baseline demographic and behavioral factors were derived from standard questionnaires for both cohorts and evaluated as risk factors for progression: age (55 to 64, 65 to 74, ≥ 75), sex, education (≤ high school, > high school), body mass index (BMI) (<25, 25 to 29, ≥ 30), and smoking status (never, past, current). Only whites were analyzed due to small numbers and low prevalence of AMD in other ethnic groups. Signs of AMD were graded based on review of color photographs and detailed phenotype information was used to classify eyes in AREDS. For SLCS, classification of AMD status was based on ocular records and fundus color photographs, as well as autofluorescent photographs and optical coherence tomography when available. Grades were assigned according to the Clinical Age-Related Maculopathy Grading System as no AMD, early, intermediate, or advanced AMD with GA or NV using the same definitions for stages of AMD for both cohorts.^1,5,23,24^ Genetic variants were identified using the same genotyping and sequencing platforms for both cohorts with quality control analyses as previously described. ^1,5,8,13,15^

### STATISTICAL ANALYSES

Associations between person-specific characteristics and type 1 *CFI* carrier status were assessed using Chi square analyses or Fisher’s exact test. For ordered categorical variables, eg. BMI, Chi square test of trend was performed. For other characteristics, eg. smoking, Chi square tests for heterogeneity were performed. We used PROC LOGISTIC of SAS and performed a stepwise analysis to identify significant genetic variants associated with progression to AAMD, with P = 0.05 for entering and staying in the model. Separate analyses were performed for progression to AAMD, GA and NV. An eye could progress to GA and then progress to NV. To assess the independent effect of the *CFI* rare variant status, we adjusted for demographic, behavioral and genetic covariates using PROC GENMOD of SAS with a logistic link and a binomial distribution and a working independence correlation structure to account for correlation between fellow eyes.

A Genetic Risk Score (GRS) was calculated from variants associated with progression to overall advanced AMD, selected from the stepwise model:

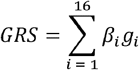

where g_i_ = number of risk alleles present for the i^th^ genetic variant I =1,…16.

Then GRS scores were grouped into tertiles. Separate GRS scores were also calculated for both GA and NV. We also evaluated the effect of *CFI* variant status according to specific GRS tertiles. The model was based on generalized estimating equation (GEE) with a binomial distribution and a logistic link. Odds ratios (OR’s) and 95% confidence intervals were calculated for progression to each outcome, with the eye as the unit of analysis, adjusted for age, sex, education, BMI, smoking, AMD baseline eye-specific grade, and study cohort. We combined the data in the two cohorts and used an indicator for study in the analysis. Using PROC PHREG, a proportional hazard regression model, we generated Kaplan-Meier survival probabilities for progression to GA by *CFI* rare variant status and by baseline AMD grade, adjusting for age, sex, education, BMI, smoking, AMD baseline eye-specific grade, GRS, *CFI* status and study cohort. Type 2 and 3 carriers were combined since very few subjects were carriers of type 2, and secondary analyses assessed the association of these variants with family history of AMD and disease progression.

## RESULTS

The characteristics of the prospective study cohorts are shown in **eTable 2 (online only)**. Among the total of 9101 eyes with non-advanced AMD at baseline, there were 90 eyes (1%) with *CFI* type 1 rare variants (53 in SLCS and 37 in AREDS). There was a greater proportion of older (>75 years) and younger (<65 years) individuals and a higher percentage of subjects with grade 1 at baseline in SLCS than in AREDS. Level of education was lower in SLCS than in AREDS, there were fewer individuals who never smoked and more past smokers in SLCS, while BMI was similar. Importantly, progression rates adjusting for age, gender, education, smoking, BMI, and AMD baseline eye-specific grade did not differ between the two cohorts for AAMD progression: comparison of hazard ratios (HR) = 0.93; 95% CI = 0.82, 1.05; P = 0.25. Rates were also similar in the two cohorts for the advanced subtypes: GA (HR = 0.89; 95% CI = 0.75, 1.06; P = 0.19) and NV (HR = 1.06, 95% CI = 0.91, 1.24; P = 0.46).

**Table 3** displays the demographic and genetic distribution of the study population based on *CFI* type 1 rare variant carrier status. Among carriers of the rare type 1 functional *CFI* variants, 44% progressed to overall AMD compared with 20% of non-carriers (P <0.001) over the 12-year follow-up period. Progression rates to advanced AMD sub-types were significantly different in *CFI* carriers as compared with non-carriers in both GA (30% vs. 10%; P<0.001) and NV (18% vs. 11%; P = 0.049). A greater percentage of type 1 carriers were grade 3 at baseline compared with non-carriers (P<0.001). A higher percentage of carriers were older than age 75 at baseline (25% versus 16%; P = 0.034). There were minimal differences for gender, education, and BMI. For smoking, 75% of carriers were ever smokers versus 56% for non-carriers (P = 0.013 for heterogeneity).

**Table 3.**
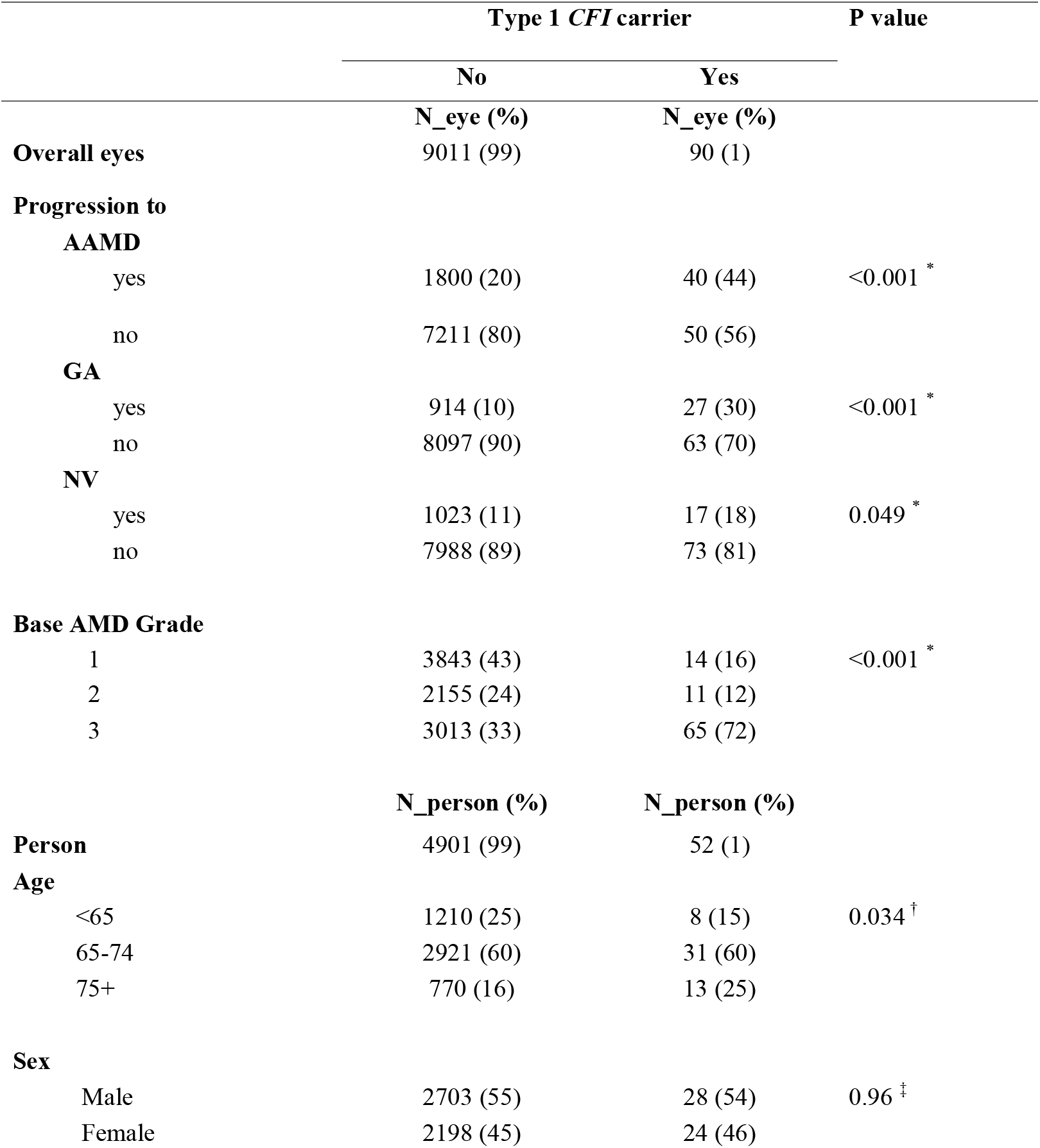
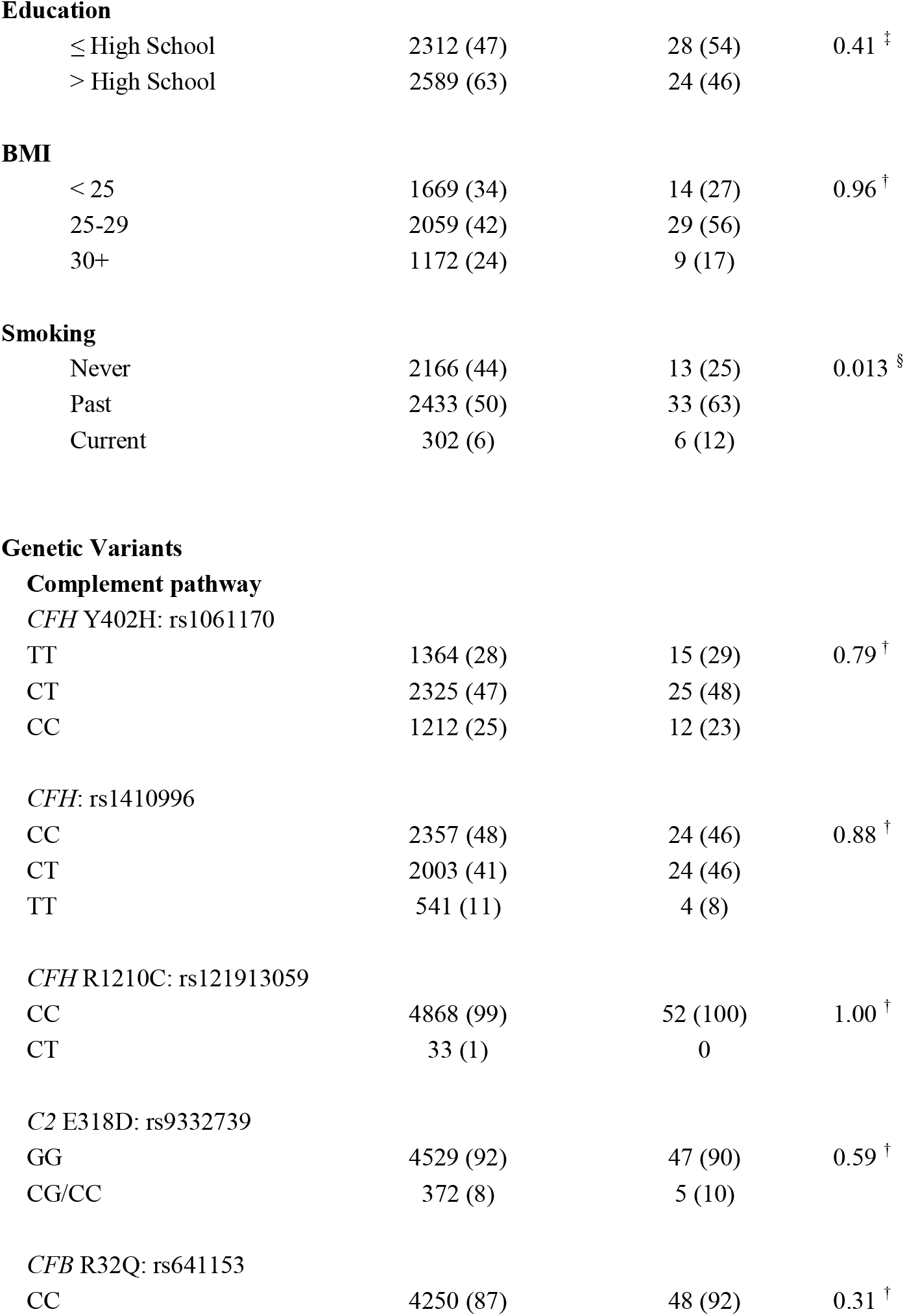
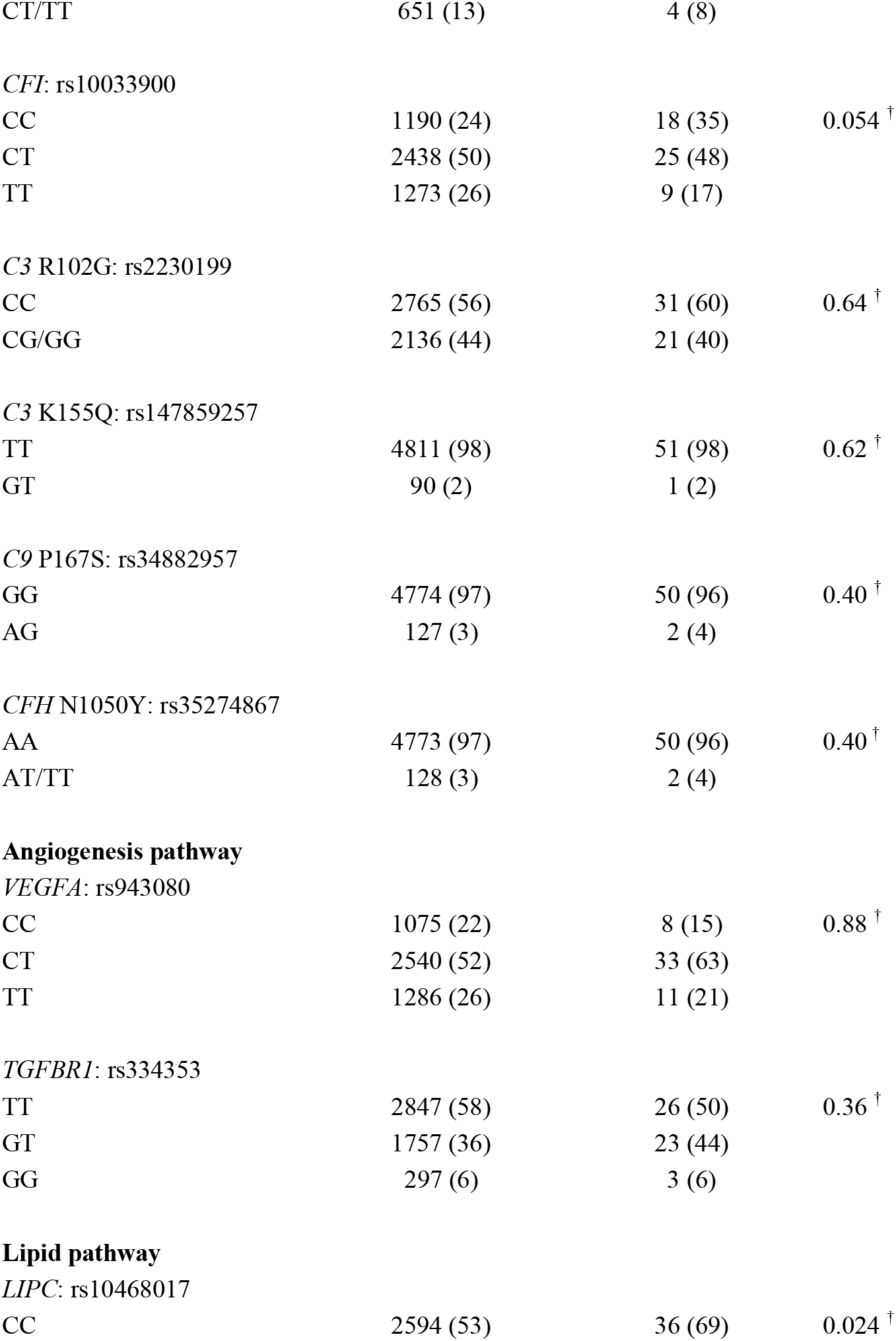
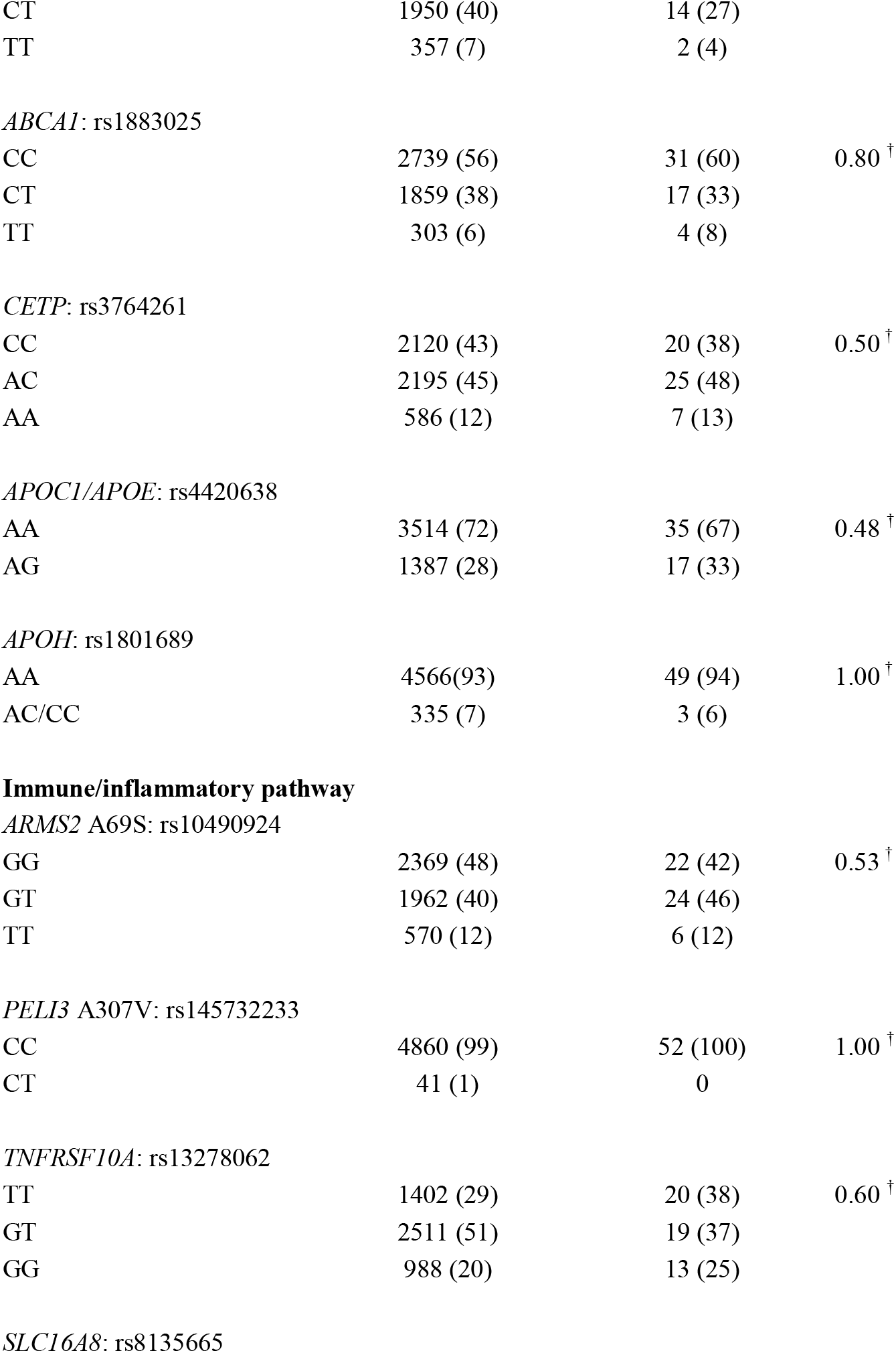
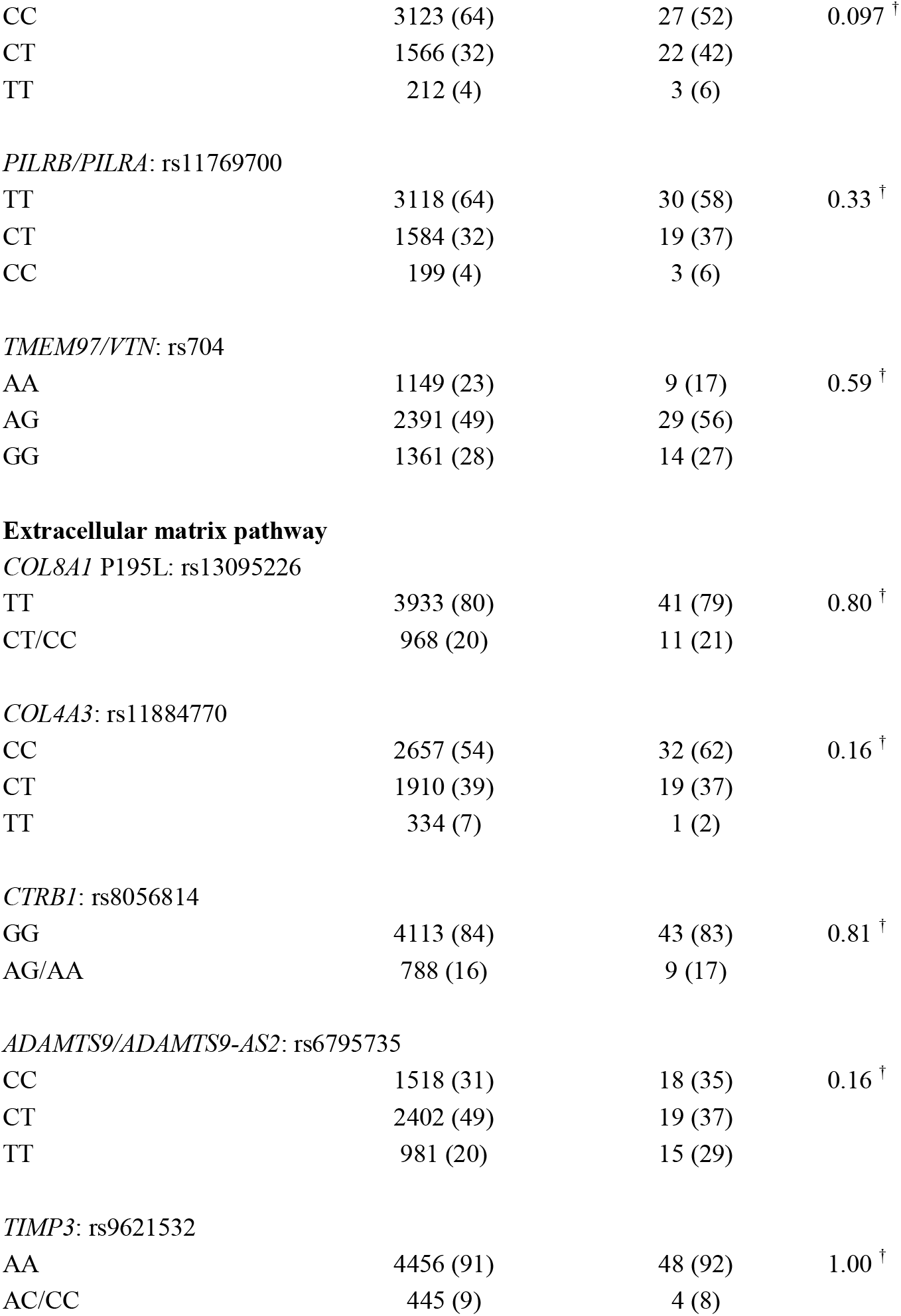
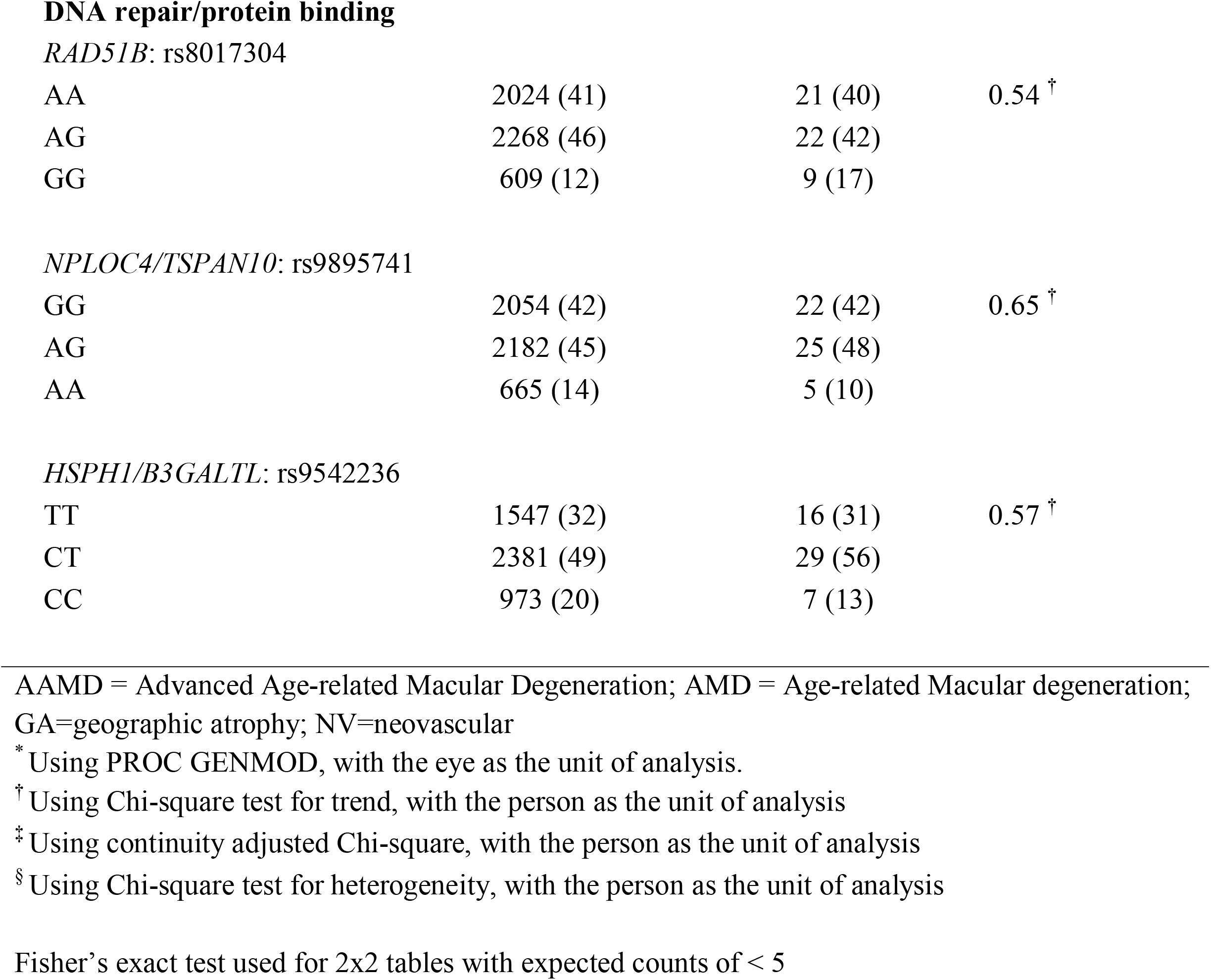
Characteristics of study population according to type 1 *CFI* rare variant carrier status.

The distributions of genotypes for most known AMD genes were similar for carriers and non-carriers. However, type 1 rare *CFI* carriers were somewhat less likely to be homozygous for the common *CFI* risk variant (P = 0.054) and the protective common *LIPC* variant ^26^ (P = 0.024). There were no significant interactions between *CFI* rare variant status and these 2 variants regarding progression to the 3 AMD outcomes.

Type 1 *CFI* rare variant status was associated with reported family history of AMD in the AREDS cohort **(Table 4**). Among 22 carriers of rare variants, 36% had 1 family member affected and 14% had 2 or more family members affected. Among subjects with no *CFI* variant, 19% had 1 family member affected and 8% had 2 or more family members affected (P for trend for *CFI* carrier vs non-carrier = 0.035). Similar family history data was not yet available in the SLCS dataset.

**Table 4.**
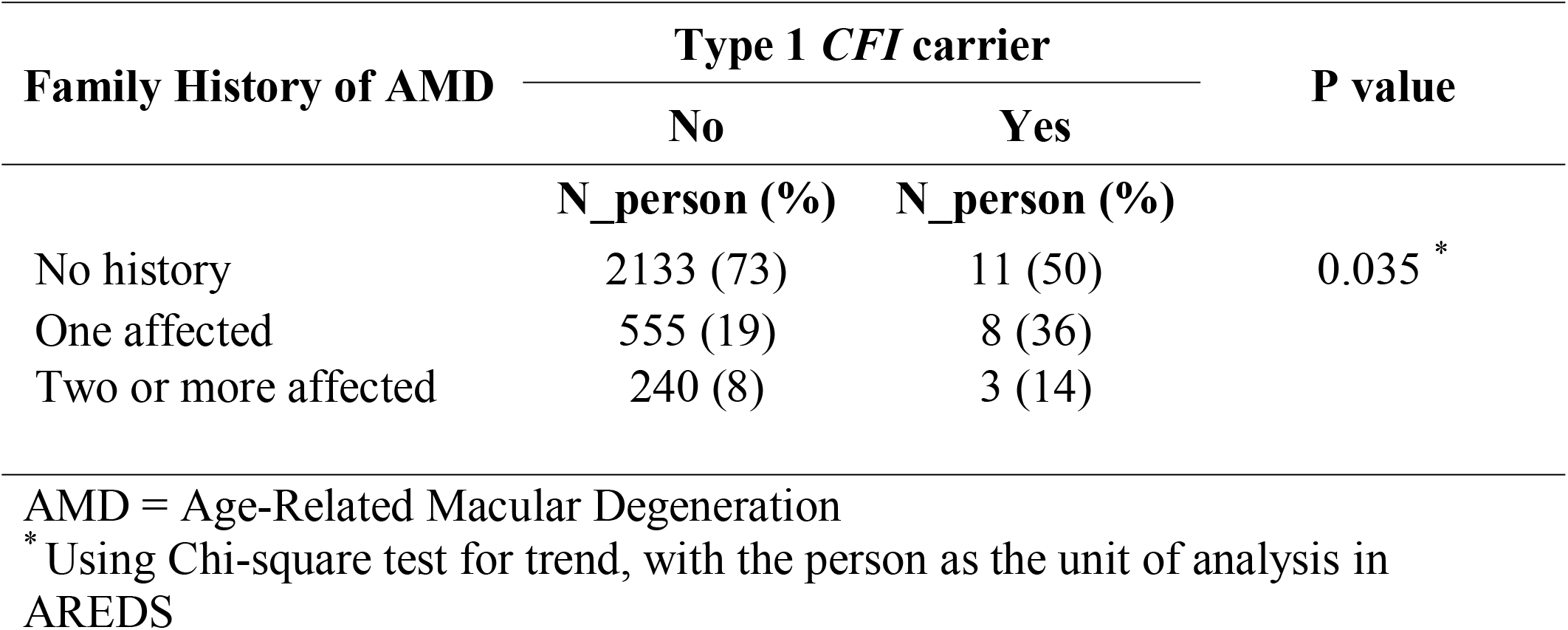
Association between type 1 *CFI* rare variant carrier status and family history of AMD.

**Table 5** shows the stepwise selection of genetic variants associated with each outcome. Nine variants in the complement pathway, one in the lipid pathway, 2 in the immune/inflammatory, 2 in the extracellular matrix, and 2 in the DNA repair/protein binding pathways were associated with progression to overall advanced AMD. Nine variants were associated with increased risk; *CFH* Y402H: rs1061170, *CFH* R1210C: rs121913059, *CFI*: rs10033900, *C3* R102G: rs2230199, *C3* K155Q: rs147859257, *C9* P167S: rs34882957, *ARMS2* A69S: rs10490924, *COL8A1* P195L: rs13095226, *HSPH1/B3GALTL*: rs9542236. Seven variants were protective; *CFH*: rs1410996, *C2* E318D: rs9332739, *CFB* R32Q: rs641153, *LIPC*: rs10468017, *TMEM97/VTN*: rs704, *RAD51B*: rs8017304, *CTRB1*: rs8056814. The highest risks for overall AMD were associated with the rare variants *CFH* R1210C (OR = 3.53; P < 0.0001) and *C3* K155Q (OR = 2.07; P = 0.0007). Regarding the advanced AMD subtypes, eight variants were related to progression to GA. Fifteen variants were related to progression to NV including some genes not related to GA in the angiogenesis and DNA repair/protein binding pathways. There were more genes in the extracellular matrix pathway associated with progression to NV than to GA. Genetic variants in the complement pathway were associated with both outcomes. *ARMS2* was associated with both outcomes with somewhat higher odds ratio for NV (OR = 1.79; P < 0.0001) than GA (OR = 1.43; P < 0.0001), consistent with previous studies. ^27^

**Table 5.**
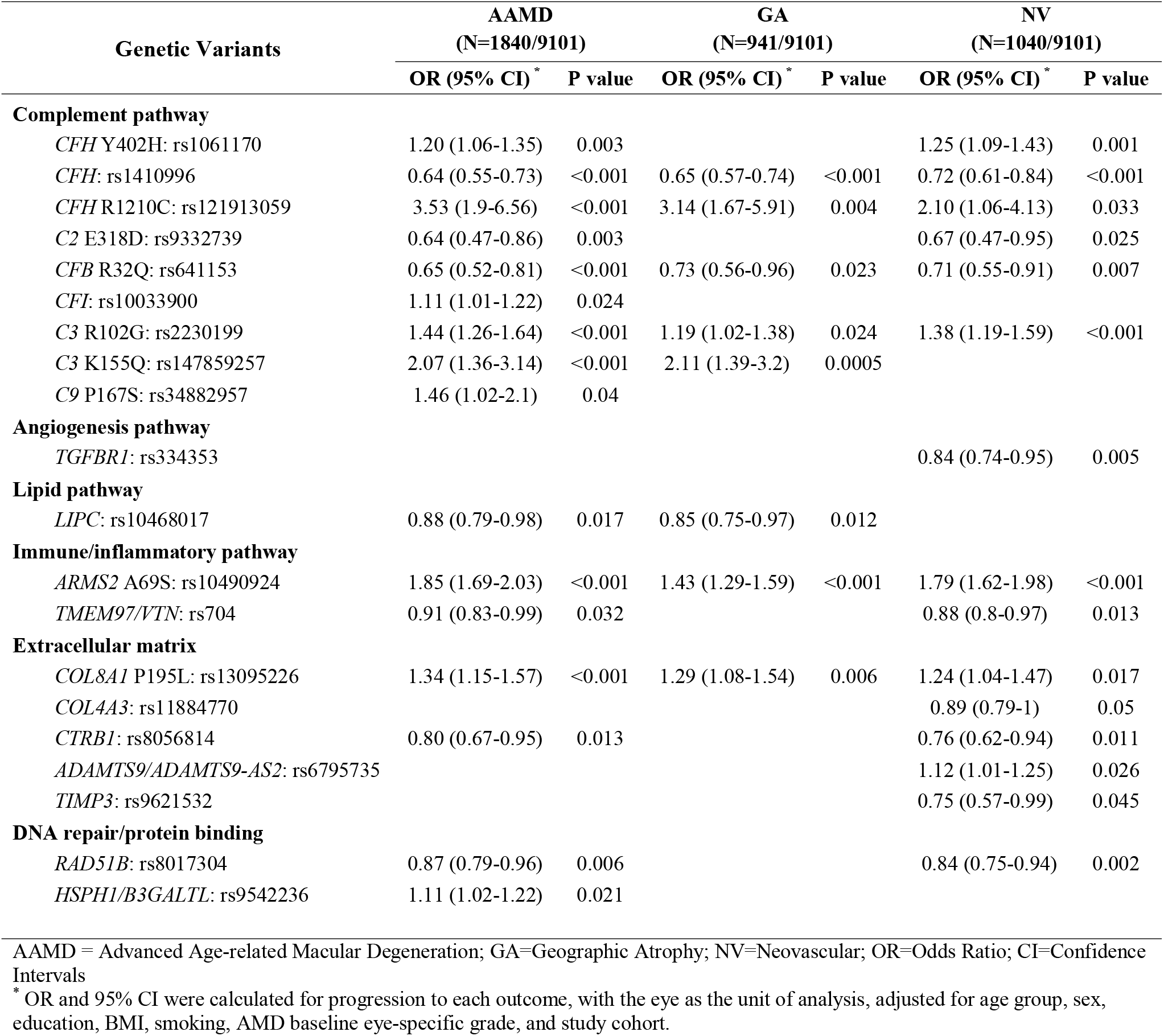
Stepwise selection of genetic variants associated with progression to AAMD, GA, and NV.

**Table 6** displays the full multivariate model to determine if type 1 *CFI* rare variant carrier status contributed to risk of progression independently of other predictors of progression. Type 1 *CFI* rare variant status was associated with a non-significant higher risk of progression to AAMD (OR = 1.49; 95% CI = 0.88, 2.53; P = 0.14). There was a modest statistically significant increased risk of progression to GA (OR = 1.91; 95% CI = 1.03, 3.52; P = 0.04), but no association with NV (OR = 0.96; 95% CI = 0.54, 1.71; P = 0.88) (**Figure 2**). As in previous analyses, age, education, higher BMI, smoking status, and baseline grade were associated with progression to AAMD, For the AAMD subtypes, education was not related to progression to GA and smoking was somewhat more strongly associated with progression to NV than GA. Many genetic variants were associated with both GA and NV. However, some genetic variants were only associated with GA (*C3* K155Q: rs147859257, *LIPC*: rs10468017), while others were only associated with NV (*CFH* Y402H: rs1061170, *C2* E318D: rs9332739, *TGFBR1*: rs334353, *TMEM97/VTN*: rs704, *COL4A3*: rs11884770, *CTRB1*: rs8056814, *ADAMTS9/ADAMTS9-AS2*: rs6795735, *TIMP3*: rs9621532). There was a higher progression rate to GA in those with type 1 *CFI* rare variant status compared to those without *CFI* rare variant status particularly with higher AMD grade at baseline, adjusting for all other covariates (**Figure 3)**.

**Table 6.**
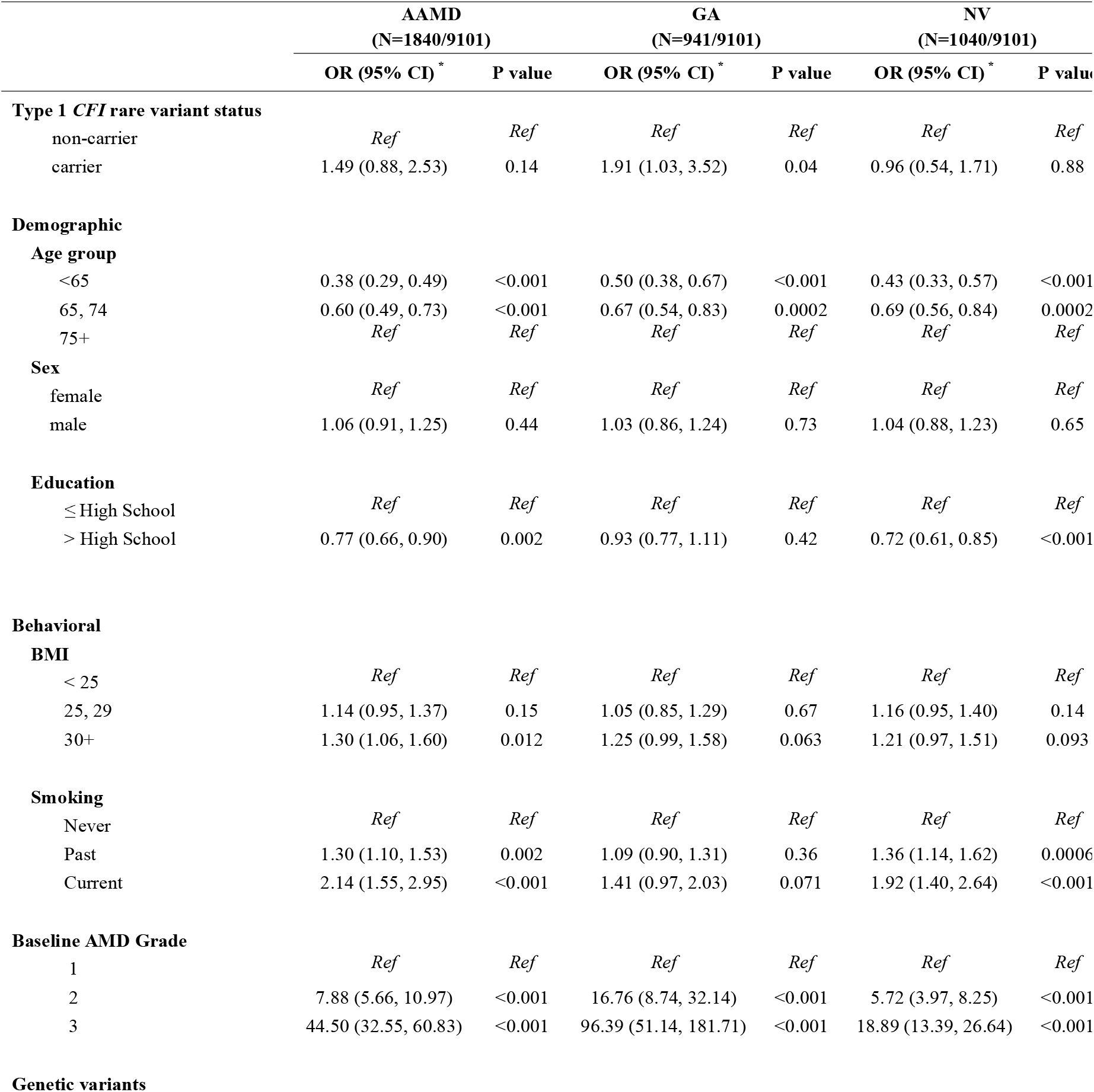
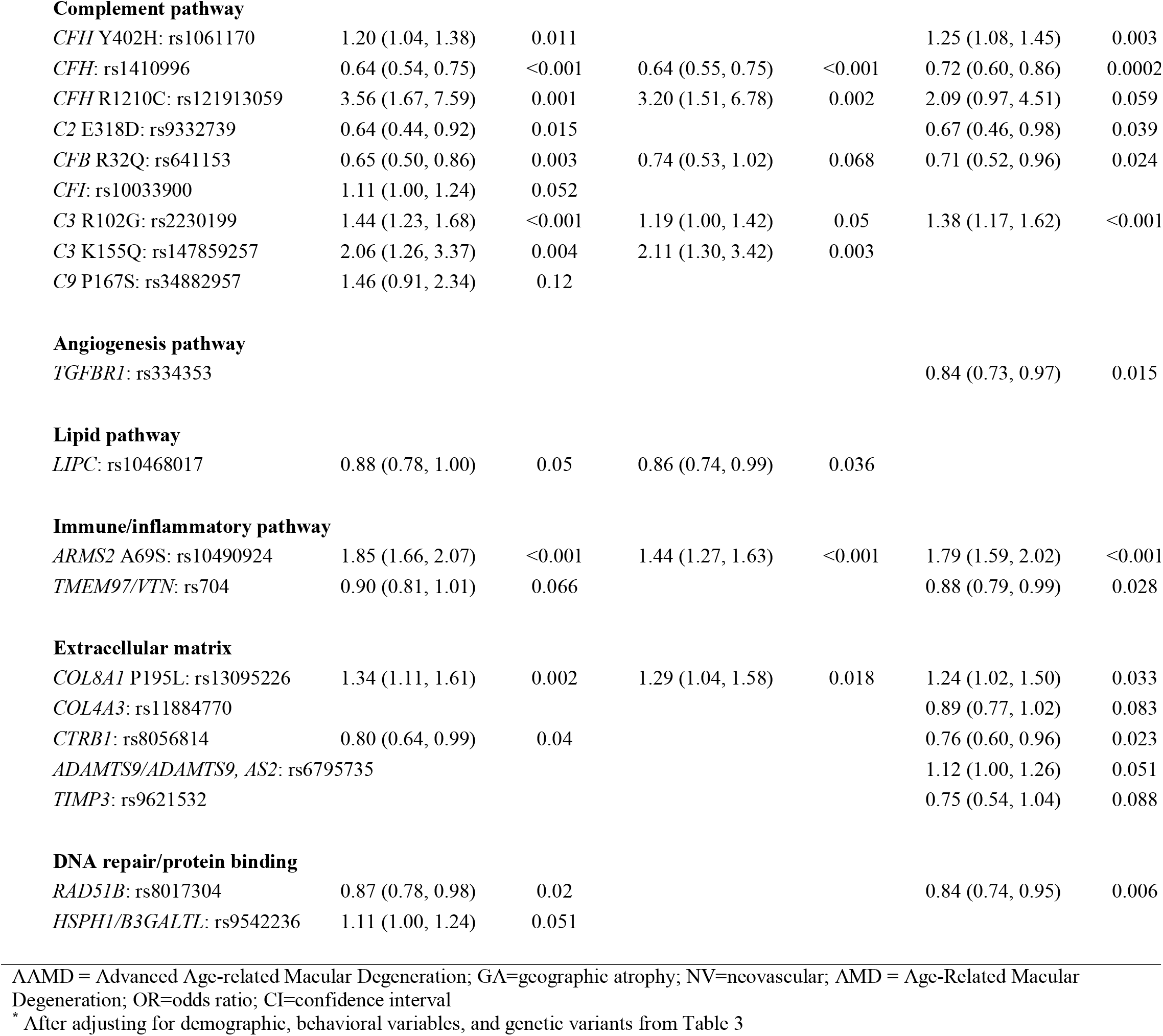
Multivariate association of type 1 *CFI* rare variant carrier status with progression to AAMD, GA, and NV.

Higher GRS values (tertiles 2 and 3) were significantly related to progression to advanced disease, compared to tertile 1, and the trend for increasing risk for higher GRS was significant for progression to AAMD, GA, and NV (P <0.001 for GRS tertile 3 vs tertile 1 for all outcomes) (**Table 7**). We evaluated whether there was an effect of type 1 *CFI* rare variant carrier status on AMD outcomes within specific GRS tertiles. There was a suggestive increased risk associated with type 1 *CFI* rare variant status among individuals with the lowest GRS tertile for progression to GA (OR = 3.65; 95% CI = 0.98,13.6; P = 0.054), meaning that type 1 *CFI* rare variant carriers in the lowest GRS had higher risk than non-carriers who are in the lowest GRS tertile. There was a non-significant trend toward a positive effect of type 1 *CFI* rare variant status on higher rate of progression to GA for all three GRS tertiles, so no significant difference in risk conferred by type 1 *CFI* rare variant status according to GRS was observed. For progression to NV, the risk of progression among type 1 *CFI* carriers was not significantly different according to GRS tertiles. Tests for the effects of interactions between GRS tertile and type 1 *CFI* rare variant status on the AMD outcomes were not significant.

**Table 7.** Effect of Genetic Risk Score (GRS) and type 1 *CFI* rare variant carrier status within categories of GRS on progression to AAMD.

|  | AAMD * |  | GA * |  | NV * |  |
| --- | --- | --- | --- | --- | --- | --- |
|  | OR (95% CI) | P value | OR (95% CI) | P value | OR (95% CI) | P value |
| GRS tertile 1 <sup>†</sup> | *Ref* |  | *Ref* |  | *Ref* |  |
| GRS tertile 2 | 2.00 (1.59-2.52) | <0.001 | 1.42 (1.06-1.90) | 0.017 | 1.72 (1.32-2.26) | <0.001 |
| GRS tertile 3 | 4.55 (3.66-5.66) | <0.001 | 2.52 (1.93-3.29) | <0.001 | 3.87 (3.00-4.97) | <0.001 |
| *CFI* type 1 carrier <sup>‡</sup> |  |  |  |  |  |  |
| GRS tertile1 | 1.46 (0.35-5.98) | 0.60 | 3.65 (0.98-13.64) | 0.054 | 1.10 (0.17-7.28) | 0.92 |
| GRS tertile2 | 1.22 (0.55-2.67) | 0.63 | 1.66 (0.63-4.39) | 0.30 | 1.01 (0.33-3.09) | 0.99 |
| GRS tertile3 | 1.69 (0.75-3.79) | 0.21 | 1.66 (0.66-4.15) | 0.28 | 0.77 (0.37-1.62) | 0.49 |

AAMD = Advanced Age-Related Macular degeneration; GRS= Genetic Risk Score; OR=Odds ratio; CI= Confidence Intervals; GA=Geographic Atrophy; NV=Neovascular; *Ref* = Reference group
\* Each model was based on GEE with a binomial distribution and a logistic link. OR and 95% CI were calculated for progression to each outcome, with the eye as the unit of analysis, adjusted for age group, sex, education, BMI, smoking, AMD baseline eye-specific grade, and study cohort.
<sup>†</sup> Each GRS was based on genes selected in Table 3 for each outcome. GRS to AAMD has a median of 0.27 (IQR: -0.10 to 0.63), and 0.00 (IQR: -0.36 to 0.35) for GA, and 0.03 (IQR: -0.43 to 0.47) for NV.
<sup>‡</sup> Effect of type 1 *CFI* carrier status within specific GRS tertiles.

**Figure 2.**
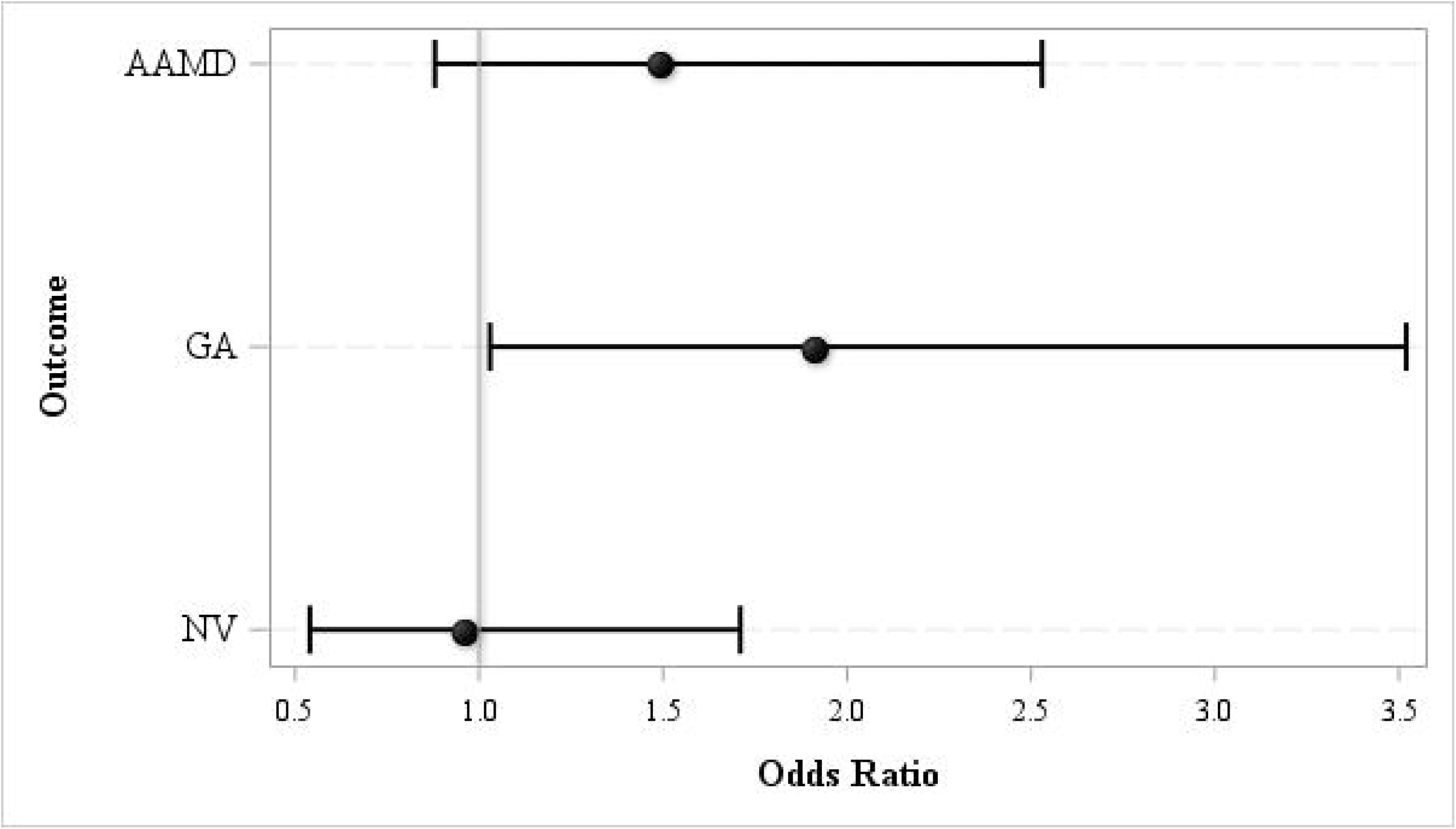
Association of type 1 *CFI* rare variant carrier status with progression to advanced AMD (AAMD), geographic atrophy (GA), and neovascular (NV), adjusting for all demographic, behavioral, and other genetic factors. Black dot = Odds Ratio; bracket line = 95% Confidence Intervals

**Figure 3.**
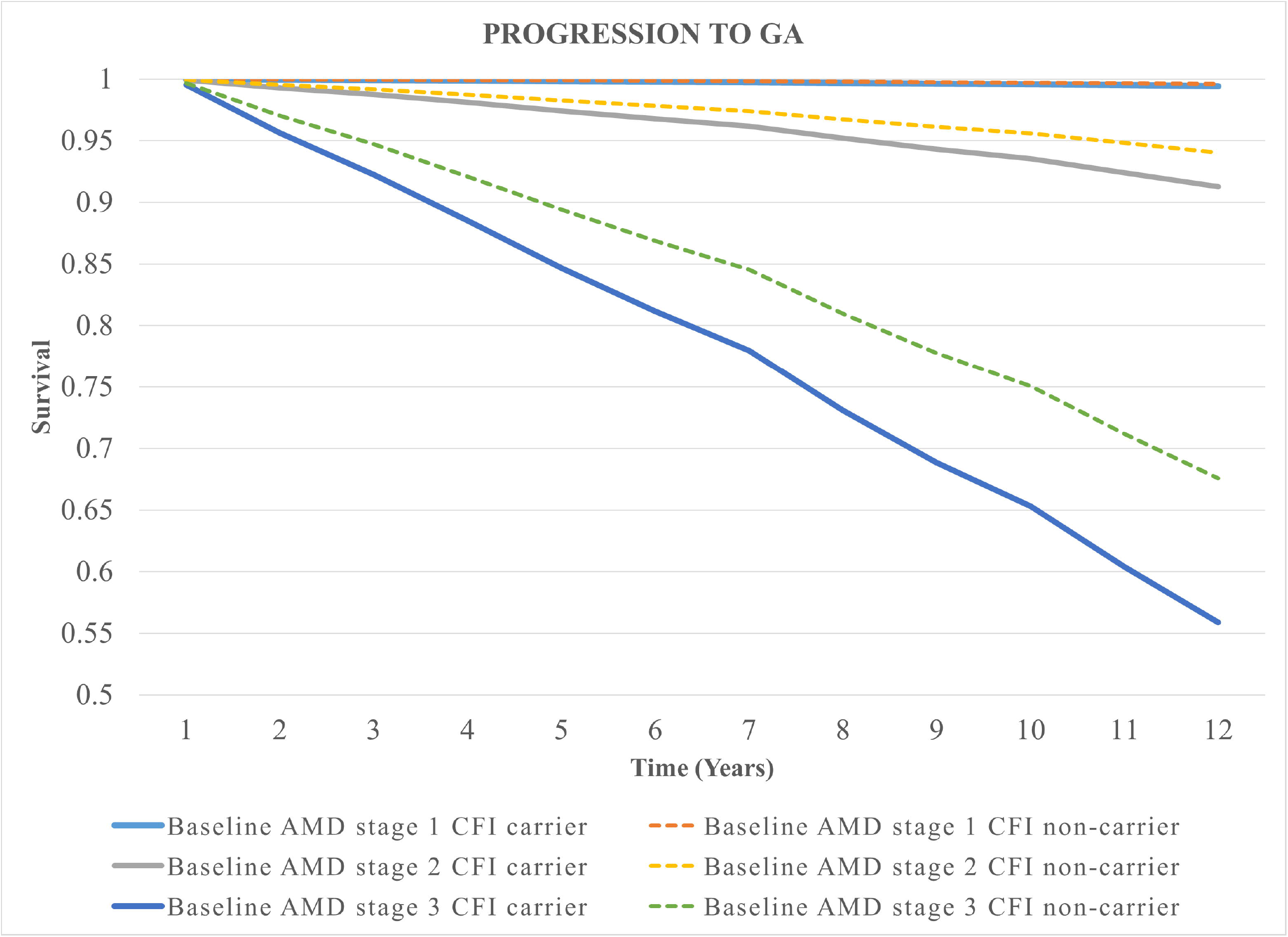
Kaplan-Meier Survival Curves for progression to geographic atrophy (GA) over 12 years by baseline AMD stages and type 1 *CFI* rare variant carrier status, adjusting for age group, sex, education, BMI, smoking, AMD baseline eye-specific grade, GRS, *CFI* status and study cohort.

We evaluated whether there were interactions between type 1 *CFI* carrier status and individual AMD genes on risk of progression. No significant interactions between type 1 *CFI* carrier status and other genetic variants were noted for progression to AAMD, GA, or NV.

As shown in **Table 8**, we conducted analyses of interactions between healthy lifestyle factors (BMI and smoking) and type 1 *CFI* carrier status to further evaluate the role of these modifiable behaviors on progression to GA and NV. The BMI-*CFI* carrier status interaction was significant for GA (P = 0.011). High BMI (≥25) was associated with a higher risk of progression to GA compared to low BMI among type 1 *CFI* carriers (OR = 5.8; 95% CI =1.5, 22.3, P = 0.011). In contrast, higher BMI ≥25 was not associated with increased rate of progression to GA among non-carriers (OR = 1.1; 95% CI = 0.9,1.3; P = 0.4). For progression to NV, a similar effect of BMI for type 1 carriers versus non-carriers was not seen: the effect of BMI among carriers was not significant (OR = 0.6; 95% CI = 0.2, 1.9; P = 0.33), whereas it was borderline significant among non-carriers (OR = 1.2; 95% CI = 1.0, 1.4; P = 0.05). Type 1 *CFI* carrier status tended to be more strongly associated with progression to GA among non-smokers (OR = 2.4; 95% CI = 0.9, 6.0; P = 0.069), adjusting for all covariates including GRS, but the interaction between smoking and *CFI* carrier status was not significant. There was no significant interaction between smoking and *CFI* carrier status for progression to NV.

**Table 8.** Association and interaction of type 1 *CFI* rare variant carrier status with lifestyle factors.

| Outcome | Exposure |  | *CFI* non-carrier |  |  | Type 1 *CFI* carrier |  |  |
| --- | --- | --- | --- | --- | --- | --- | --- | --- |
|  |  |  | OR | 95% CI | P value | OR | 95% CI | P value |
| AAMD | BMI<sup>*</sup> | < 25 | 1.0 (*Ref*) | - | - | 0.7 | (0.3, 2.0) | 0.55 |
|  |  | ≥ 25 | 1.2 | (1.0, 1.4) | 0.049 | 2.1 | (1.2, 3.9) | 0.016 |
| GA |  | < 25 | 1.0 (*Ref*) | - | - | 0.5 | (0.2, 1.6) | 0.24 |
|  |  | ≥ 25 | 1.1 | (0.9, 1.3) | 0.40 | 3.0 | (1.5, 6.1) | 0.002 |
|  | BMI ≥25 vs <25 |  | 1.1 | (0.9, 1.3) | 0.40 | 5.8 | (1.5, 22.3) | 0.011 |
| NV |  | < 25 | 1.0 (*Ref*) | - | - | 1.4 | (0.6, 3.4) | 0.48 |
|  |  | ≥ 25 | 1.2 | (1.0, 1.4) | 0.054 | 0.9 | (0.4, 1.2) | 0.69 |
|  | BMI ≥25 vs <25 |  | 1.2 | (1.0, 1.4) | 0.054 | 0.6 | (0.2, 1.9) | 0.33 |
| AAMD | Smoking<sup>†</sup> | Never | 1.0 (*Ref*) | - | - | 2.0 | (0.7, 5.5) | 0.21 |
|  |  | Ever | 1.3 | (1.1, 1.6) | < 0.001 | 1.7 | (0.9, 3.2) | 0.10 |
| GA |  | Never | 1.0 (*Ref*) | - | - | 2.4 | (0.9, 6.0) | 0.069 |
|  |  | Ever | 1.1 | (0.9, 1.3) | 0.31 | 1.8 | (0.8, 4.0) | 0.14 |
| NV |  | Never | 1.0 (*Ref*) | - | - | 1.2 | (0.4, 3.2) | 0.73 |
|  |  | Ever | 1.4 | (1.2, 1.7) | < 0.001 | 1.1 | (0.5, 2.3) | 0.88 |

AAMD = Advanced Age-related Macular Degeneration; GA=Geographic Atrophy; NV=Neovascular; OR = Odds Ratio; CI = Confidence Intervals; *Ref* = Reference Group
<sup>\*</sup> Adjusted for age (categories), sex, education, ever/never smoking, AMD baseline–eye specific grade, genetic risk score (tertiles) and study cohort
<sup>†</sup> Adjusted for age (categories), sex, education, BMI (≥ 25, < 25), AMD baseline – eye specific grade, genetic risk score (tertiles) and study cohort

A secondary analysis assessed type 2 (N= 5 subjects, 9 eyes) and type 3 variants (N= 106 subjects, 193 eyes), and due to the small number of type 2 subjects these were combined (N = 111 subjects, 202 eyes). Type 2-3 *CFI* rare variant status was not associated with reported family history of AMD in the AREDS cohort (P for trend = 0.69). In the total cohort (SLCS and AREDS), after adjusting for genes selected in the stepwise regression, *CFI* type 2-3 rare variant status was not associated with risk of progression to AAMD (OR = 0.98; 95% CI = 0.55, 1.89; P = 0.94). Similar non-significant results were seen with progression to GA and NV.

## DISCUSSION

We report new information indicating that carriers of type 1 *CFI* rare genetic variants with low serum antigenic levels and reduced function are at higher risk of progression from non-advanced AMD to AAMD with GA. *CFI* dysfunctional rare variant carriers had a statistically significant 1.9-fold increased risk of progression to GA. After adjusting for all other covariates, there was no association between *CFI* carrier status and risk of progression to NV. Carriers of the rare type 1 *CFI* variants were also more likely to have other family members with AMD, which expands previous observations of complement rare variants occurring in families with AMD. ^8–12^ Similar effects were not seen for less dysfunctional rare *CFI* variants.

We evaluated the effect of behavioral risk factors according to *CFI* carrier status and found that high BMI was associated with a higher risk of progression to GA compared to low BMI, among *CFI* carriers but not among non-carriers. The analyses according to smoking were not significant. Results imply that behavioral modifiable factors may play a role to some extent in the complex genetic susceptibility conferred by rare variants with high impact. We and others have previously shown that BMI and smoking add to the effect of the more common genetic variants on risk of progression to AAMD. ^1,5,6^

Complement activation is designed to be devastating to a pathogen but, if misdirected or dysregulated, it can be similarly damaging to self-tissue.^18,19^ Control of the complement system is therefore mediated by both plasma and cell-bound regulators. Factor I is a serine protease in the blood that modulates the complement cascade through proteolytic cleavage of complement component C3b, in conjunction with cofactor proteins such as Factor H or Membrane Cofactor Protein (MCP, CD46). Loss of function of a complement regulator, such as a rare genetic variant in *CFI*, can thus lead to an inadequately regulated complement cascade. We have previously conducted serum-based functional studies for rare *CFI* variants in AMD and demonstrated that type 1 variants lead to low serum FI antigenic levels and a corresponding decrease in FI function.^14,22^ The existing hypothesis is that the dysfunctional genetic *CFI* variants, in the presence of a trigger or a given degree of retinal injury, predispose to excessive complement activation that accelerates damage to the retina. Our current results support the association between dysfunctional *CFI* rare variants (particularly Type 1 variants that lead to half-normal FI antigenic levels) and long-term AMD outcomes.

Compared to common variations, the rare mutations have large effects and clearer biologic consequences. Discovery of rare variants opened a new line of inquiry and research for AMD pathogenesis and treatment. For example, identification of rare variants in *CFH* ^8–13^ supported targeting the alternative complement pathway activity components and/or restoring the levels of this complement inhibitor to normal levels (NCT04643886). The discovery of the association between AMD and the *C3* common and rare variants^13,28^ supported the development and assessment of therapeutic targets aimed at C3 for GA (NCT03525613 and NCT03525600). As is the case for Factor H protein, therapeutic agents focusing on FI protein are in clinical trials for treatment of AMD (NCT03846193, NCT04566445, NCT04437368). Our new results indicate that type I rare *CFI* variants increase the risk of progression to GA, providing additional clinical evidence and support for these approaches.

These types of *CFI* rare variants have also been described to be causative in 5-15% of patients with a kidney disease, called atypical hemolytic uremic syndrome (aHUS).^29^ Hemolytic uremic syndrome is a thrombotic microangiopathy that features acute endothelial injury and often manifests in early childhood or young adults, whereas AMD is a disease primarily in older adults. Given the striking overlap of ∼40-50% variants causing two apparently disparate diseases and a possible association between late AMD and reduced kidney function,^14,30^ we searched the databases in our SLCS cohort for potential associations. We selected in advance only those diagnoses potentially relevant to the complement pathway including glomerulonephritis, end stage kidney disease due to preeclampsia and genetic kidney disease leading to kidney failure, and then tested the association of these kidney diagnoses with the rare dysfunctional Type 1 variants. Among the carriers (N = 36), 8.3% had these diagnoses and among the non-carriers (N = 2079), 3.4% reported these kidney disorders, with an OR of 2.57, P = 0.13 (Fisher’s Exact Test). Although the numbers were small and non-significant, there was a larger percent of these diagnoses among the *CFI* carriers.

Strengths of the current study include the standardized assessment of risk factors and AMD outcomes in both cohorts, the longitudinal follow-up of comparable duration in both groups, and similar rates of progression adjusting for baseline covariates. We conducted analyses according to “eyes” since the outcome of progression to advanced AMD is heterogeneous and may differ in both type of advanced AMD and rate of progression between fellow eyes. However, outcomes for fellow eyes are not independent; therefore, our analyses specifically accounted for inter-eye correlation. Rare variant status was assessed using the same genotyping arrays and sequencing for both cohorts.

Limitations of our study include the low prevalence of the rare variants requiring the combination of cohorts. The mechanisms whereby *CFI* variants were associated with progression to GA and not NV in these analyses is unclear, and further research is warranted to confirm and expand upon these findings. Replication in another large prospective cohort would be helpful to confirm these findings.

In summary, we present new results pointing out that rare, dysfunctional variants in the *CFI* gene are associated with family history of AMD and progression to the advanced stage of AMD with geographic atrophy, which can lead to significant loss of vision and has no currently approved therapy. Prediction modeling should incorporate both common and rare genetic variants to identify those at highest risk of progression to advanced stages of AMD. These results support the identification of carriers of rare variants with high impact in order to target specific treatments to susceptible individuals. ^5,6,31^

## Supporting information

eTable 1

eTable 2

## Data Availability

Data for collaborative studies are available upon reasonable request to the authors

## Abbreviations

(*CFI*): Complement Factor I
(FI): Factor I protein
(AMD): Age-Related Macular Degeneration
(AAMD): Advanced Age-Related Macular Degeneration
(GA): Geographic Atrophy
(NV): Neovascular
(SLCS): Seddon Longitudinal Cohort Study
(AREDS): Age-Related Eye Disease Studies
(BMI): Body Mass Index
(*CFH*): Complement Factor H
(*C3)*: Complement Component 3
(*C9)*: Complement Component 9
(FH): Factor H
(C4BP): C4 Binding Protein
(MCP; CD46): membrane cofactor protein
(CR1; CD35): complement receptor 1
(SAS): Statistical Analysis System
(GRS): Genetic Risk Score
(HR’s): Hazard Ratio’s
(GEE): Generalized Estimating Equation
(OR’s): Odds Ratio’s
(aHUS): Atypical Hemolytic Uremic Syndrome

