## Supplementary material for "Rare Dysfunctional Complement Factor I Genetic Variants and Progression to Advanced Age-Related Macular Degeneration": eTable 1

**eTable 1. List of three types of *CFI* genetic variants.**

| ***Type I*** | ***Type II*** | ***Type III*** |
|---|---|---|
| A240G | A356P | C54* |
| A258T | E554V | D403N |
| C467R | G362A | G261D |
| D310E | G487C | I492L |
| E109A | G500R | K441R |
| G119R | N536K | P553S |
| G162D | R339Q | Q462H |
| G263V | V184M | R317W |
| G287R | V20I | R406H |
| H418L | | T300A |
| IVS11+1G>C | | |
| N177I | | |
| P50A | | |
| P64L | | |
| Q217H | | |
| R202I | | |
| R474* | | |
| R502C | | |
| S221Y | | |
| V152M | | |
| V230M | | |
| V543A | | |
| W541* | | |
