## Supplementary material for "Rare Dysfunctional Complement Factor I Genetic Variants and Progression to Advanced Age-Related Macular Degeneration": eTable 2

**eTable 2. Characteristics of study cohorts**

| | Study | | |
|---|---|---|---|
| | **AREDS** | **SLCS** | **P value** |
| | **N_eye (%)** | **N_eye (%)** | |
| **Total eyes** | 5200 (57) | 3901 (43) | |
| **Type 1 *CFI* Carrier** | 37 (0.7) | 53 (1.4) | |
| **Progression to AAMD** | 13/37 (35) | 27/53 (51) | |
| **Progression to GA** | 8/37 (22) | 19/53 (36) | |
| **Progression to NV** | 8/37 (22) | 9/53 (17) | |
| **Non-Carriers** | 5163 | 3848 | |
| **Progression to AAMD** | 1142/5163 (22) | 658/3848 (17) | |
| **Progression to GA** | 576/5163 (11) | 338/3848 (9) | |
| **Progression to NV** | 669/5163 (13) | 354/3848 (9) | |
| **Progression to** | | | |
| **AAMD** | | | |
| yes | 1155 (22) | 685 (18) | HR 0.93, P=0.25* |
| no | 4045 (78) | 3216 (82) | |
| **GA** | | | |
| yes | 584 (11) | 357 (9) | HR 0.89, P=0.19* |
| no | 4616 (89) | 3544 (91) | |
| **NV** | | | |
| yes | 677 (13) | 363 (9) | HR 1.06, P=0.46* |
| no | 4523 (87) | 3538 (91) | |
| **Baseline AMD Grade** | | | |
| 1 | 1956 (38) | 1901 (49) | <0.001† |
| 2 | 1292 (25) | 874 (22) | |
| 3 | 1952 (38) | 1126 (29) | |
| | **N_person (%)** | **N_person (%)** | |
| **Person** | 2837 | 2116 | |
| **Type 1 *CFI* Carrier** | 22 (0.8) | 31 (1.5) | |
| **Age of study cohorts** | | | |
| <65 | 652(23) | 566 (27) | 0.026‡ |

| | | | |
|---|---|---|---|
| 65-74 | 1831 (65) | 1121 (53) | |
| 75+ | 354 (12) | 429 (20) | |
| Mean Age (SD) | 68.8 (4.9) | 68.9 (6.6) | 0.86‡ |
| Range | 55-81 | 55-80 | |
| **Sex** | | | |
| Male | 1249 (44) | 973 (46) | 0.18‡ |
| Female | 1588 (56) | 1143 (54) | |
| **Education** | | | |
| ≤ High School | 953 (34) | 1387 (66) | <0.001‡ |
| > High School | 1884 (66) | 729 (34) | |
| **BMI** | | | |
| < 25 | 939 (33) | 744 (35) | 0.14‡ |
| 25-29 | 1207 (43) | 881 (42) | |
| 30+ | 690 (24) | 491 (23) | |
| **Smoking** | | | |
| Never | 1336 (47) | 843 (40) | <0.001‡ |
| Past | 1332 (47) | 1134 (54) | |
| Current | 169 (6) | 139 (7) | |

SLCS = Seddon Longitudinal Cohort Study; AAMD = Advanced Age-Related Macular Degeneration; GA = Geographic Atrophy; NV = Neovascular; AMD = Age-Related Macular Degeneration

* P values for differences in Hazard Ratio's (HR's) between the studies were obtained using PHREG, adjusted for age group, sex, education, BMI, smoking, and AMD baseline eye-specific grade.

† Using PROC GENMOD, with the eye as the unit of analysis.

‡ Using Chi-square test for heterogeneity, with the person as the unit of analysis
